# UPhAIR: A Hybrid Pipeline for Generating Understandable Post-hoc AI Reports in Glioma IDH Mutation Status Prediction

**DOI:** 10.64898/2026.05.01.26349658

**Authors:** Arman Gorji, Hossein Shahverdi, Amirhossein Saberi, Benyamin Gheiji, Somayeh Farahani, Ghasem Azemi, Antonio Di Ieva

**Affiliations:** Neuroscience & Neoplasia AI research Group (NAIRG), Department of Neuroscience, Hamadan University of Medical Sciences, Hamadan, Iran; Department of Electrical and Computer Engineering, Shahid beheshti university, Tehran, Iran; Department of Immunology, School of Medicine, Iran University of Medical Sciences, Tehran, Iran; Student Research Committee, Faculty of Medicine, Mashhad University of Medical Sciences, Mashhad, Iran; Computational NeuroSurgery (CNS) Lab, Faculty of Medicine, Health and Human Sciences, Macquarie Medical School, Macquarie University, Sydney, NSW, Australia

## Abstract

Clinical adoption of machine learning (ML) in medical imaging is limited by the lack of interpretability. To address this, we present understandable post-hoc artificial intelligence reports (UPhAIR), a pipeline designed to generate transparent, evidence-based explanations by combining Shapley additive explanation (SHAP) analysis with retrieval-augmented generation (RAG) and large language models (LLMs). We trained 12 Classifiers to predict Isocitrate dehydrogenase (IDH) mutation status in glioma using radiomics and clinical features. SHAP values were used to identify key contributors to each prediction. Domain literature was collected from three sources and indexed within a RAG framework. Relevant papers were retrieved using Facebook AI similarity search (FAISS) vector similarity search and provided to Google Gemini 2.5 Pro to generate concise, reference-supported explanations for each feature. The model achieved a best AUC of 0.90±0.02 on a 5-fold cross-validation using an extreme gradient boosting (XGBoost) Classifier and a hold-out test AUC of 0.86. In a case study of a single patient excluded from training, the model correctly predicted the patient to be IDH-wildtype glioma, and SHAP identified MGMT status, age, and three radiomic features as the most influential features. UPhAIR produced a structured report combining SHAP visualizations with LLM-generated summaries grounded in scientific evidence. UPhAIR provides a practical, model-agnostic framework that enhances ML interpretability in clinical settings, helping bridge the gap between black-box AI and real-world medical decision-making.

## 1. Introduction

Artificial intelligence (AI) and machine learning (ML) are rapidly reshaping medicine, with models now capable of interpreting diagnostic images, forecasting patient outcomes, prioritizing care, and even predicting treatment responses with human-level or superior accuracy [1–3]. These advances rely on complex architectures such as convolutional neural networks, transformers, and gradient-boosting ensembles. However, their impressive performance often comes at the expense of transparency, resulting in the so-called “black-box” problem [4]. In clinical settings, where decisions carry significant consequences, practitioners and regulators demand not only accurate predictions but also clear, defensible explanations of how those predictions were made [5,6]. Without such interpretability, even the most powerful AI tools struggle to gain trust, and their adoption remains limited to research environments rather than everyday clinical practice [7].

The emerging discipline of explainable AI (XAI) directly tackles this opacity by shining light into the internal logic of sophisticated models, allowing clinicians to assess, question, and ultimately rely on AI recommendations [8]. Widely adopted, model-agnostic approaches like Shapley Additive exPlanations (SHAP) and Local Interpretable Model-Agnostic Explanations (LIME) convert complex, high-dimensional representations into numerical feature-importance scores, thereby identifying which inputs most strongly influenced a prediction [9]. However, these outputs, whether visualized as force-plots, bar charts, or extensive tables of values, still demand a level of data-science expertise that may not be available in busy clinical settings. In practice, physicians require concise, accessible explanations that seamlessly integrate with their decision-making workflows.

Large language models (LLMs) present a promising avenue for bridging this interpretability gap by converting quantitative feature attributions into coherent, clinically relevant narratives. Several studies have demonstrated that LLM-generated explanations can enhance trust and bolster decision confidence among healthcare providers and other non-technical stakeholders [10]. However, LLMs are susceptible to “hallucinations,” producing fluent but inaccurate statements [10,11]. Recent evaluations in medical contexts highlight this risk and recommend mitigation strategies including retrieval-augmented generation (RAG), rigorous prompt design, and iterative self-reflection mechanisms to ensure explanation fidelity and reliability [12,13].

In response to these challenges, we introduce UPhAIR (Understandable Post-hoc AI Reports), a versatile hybrid pipeline that combines the quantitative precision of SHAP (or any feature-attribution technique) with the expressive power of a carefully guided large language model. By employing a structured prompt design and incorporating a curated corpus of domain-specific literature using RAG, UPhAIR grounds its narrative explanations in verifiable evidence. This approach not only preserves numerical rigor but also actively mitigates the risk of LLM “hallucinations,” ensuring that every sentence can be traced to a reliable source.

We demonstrate UPhAIR’s practical utility by applying it to the well-studied task of predicting isocitrate dehydrogenase (IDH) mutation status in glioma. IDH mutation is a key biomarker in neuro-oncology integrated into the 2021 WHO central nervous system tumor classification and critically shapes patient prognosis and therapeutic decision-making [14–16]. Importantly, extensive meta-analyses covering more than 1,700 patients report pooled AUCs around 0.90 for radiomics-based IDH prediction, confirming it as a robust benchmark for evaluating new interpretability methods [17–19]. Glioma data are particularly rich, combining high-dimensional radiomic descriptors (capturing tumor morphology, texture, and intensity from MRI) with essential clinical variables such as patient age, gender, and MGMT-promoter methylation status [20]. This multi-modal dataset provides an ideal testbed for UPhAIR’s dual-format reports, which pair clear numerical feature attributions with concise, clinician-focused narrative explanations.

## 2. Methods

Our study is structured around a multi-stage pipeline designed to predict IDH mutation status in glioma and generate an explanatory report.

### UPhAIR: A Retrieval-Augmented Pipeline for Post-hoc Model Explanation

UPhAIR is a modular RAG pipeline designed to generate transparent, reference-supported explanations for machine learning model predictions in biomedical settings. Rather than providing generic justifications, UPhAIR connects model features with semantically aligned scientific literature to ground predictions in real-world evidence.

The pipeline begins by formulating a task-specific query based on the clinical prediction objective in this study, IDH mutation status prediction in glioma. This query is automatically submitted to PubMed, Semantic Scholar, and CrossRef, and all relevant open-access full-text articles are downloaded and preprocessed. References are removed, and clean, readable document text is extracted. Each document is embedded using the all-MiniLM-L6-v2 sentence transformer model to produce dense vector representations, which are stored in a Facebook AI similarity search (FAISS) index for efficient semantic retrieval.

Once a prediction is made by the trained ML model, SHAP is used to identify the most influential features driving the prediction. These features are transformed into descriptive, human-readable phrases (e.g., “tumor volume,” “age,” “enhancing component”) and used to query the FAISS index. For each top feature, the pipeline retrieves the top 3 most relevant documents, which collectively serve as a personalized literature base contextualizing the prediction.

The prompting strategy is carefully designed to ensure accurate and relevant LLM outputs. We use a structured prompt that includes: (1) a system instruction defining the LLM’s role as a biomedical expert writing a clinical justification; (2) a few-shot example to enforce output format, tone, and citation style; (3) the retrieved documents supporting the explanation; (4) the model’s prediction; and (5) the SHAP-derived key features. This prompt is fed into an LLM, which is explicitly instructed to ground its explanation only in the retrieved literature. The complete prompt is provided in Supplementary Figure 1. The result is a coherent, medically relevant report that cites specific studies to justify the AI’s prediction.

This system is fully automatic and adaptable to any binary classification model with interpretable features. The focus is on transparency and trust: each explanation is tightly linked to peer-reviewed evidence, reducing black-box opacity and offering clinicians a clear window into the rationale behind model decisions.

To demonstrate the pipeline, we applied UPhAIR to the task of IDH mutation prediction in glioma, as described in the following section.

### Showcase: IDH Mutation Prediction in Glioma

To demonstrate the utility of UPhAIR, we applied it to the clinically meaningful task of predicting IDH mutation status in glioma patients. We used publicly available T1-weighted MRI data and tumor segmentations from the University of Pennsylvania (UPenn) and University of California, San Francisco (UCSF) glioma cohorts available through The Cancer Imaging Archive (TCIA). All data used in this study are derived from publicly available, open-access datasets that have been fully anonymized prior to release. The included variables, such as patient identifiers, age, and dates, are part of the original dataset and are not linked to any personally identifiable information. These identifiers are dataset-specific and cannot be traced back to individual patients. We have retained them in their original form to ensure data integrity, reproducibility, and consistency with the source datasets. Inclusion criteria were 1) the presence of IDH mutation status, 2) the presence of T1W MRI, and 3) the presence of tumor segmentation, which resulted in a total of 1096 patients. Detailed patient demographic information is provided in Supplementary Table 1. Radiomic features were extracted from previously segmented ROI using PyRadiomics [21] with 107 standardized RFs as our reference RF set. These features encompassed 18 First-Order (FO), 15 Shape-based features (SF), 23 from the Gray Level Co-occurrence Matrix (GLCM), 16 from the Gray Level Size Zone Matrix (GLSZM), 16 from GLRLM, 5 from the Neighborhood Gray Tone Difference Matrix (NGTDM), and 14 from the Gray Level Dependence Matrix (GLDM). Clinical variables included age, sex, and MGMT methylation status. All continuous features were scaled using Min-Max normalization, and all missing features were handled using a mean imputer.

A wide range of machine learning models were evaluated for this binary classification task, including ensemble methods (Gradient Boosting, XGBoost, LightGBM, Bagging, Random Forest, AdaBoost), neural networks (MLP), distance-based models (KNeighbors), linear models (Logistic Regression), margin-based classifiers (SVC), tree-based models (Decision Tree), and probabilistic classifiers (Gaussian Naive Bayes). We optimized hyperparameters using grid search with 5-fold cross-validation on 80% of the data. Model performance was assessed on the remaining 20% test set using metrics such as AUC and F1-score. The best-performing model was then used to generate patient-level SHAP values using the kernel shap method.

These SHAP values were passed to UPhAIR as input for report generation. Each feature with high importance was matched to relevant literature using the FAISS index. The retrieved context, together with the prediction and an appropriate prompt structure, was fed into Gemini 2.5 Pro to generate personalized, citation-supported narratives explaining each prediction.

This case study demonstrates how UPhAIR can translate black-box predictions into transparent, literature-grounded explanations, helping bridge the gap between AI models and clinical trust.

## 3. Results

### 3.1. Prediction Model Performance

The model selection process identified the Boost classifier as the top-performing model. On the 80% 5FVC, it achieved an AUC of 0.90±0.02 and 0.86 on 20% hold-out test, demonstrating a strong capability to distinguish between IDH-mutant and IDH-wild-type gliomas. Detailed performance metrics and model hyperparameters are provided in Supplementary Table 2 and 3.

**Table 1.**
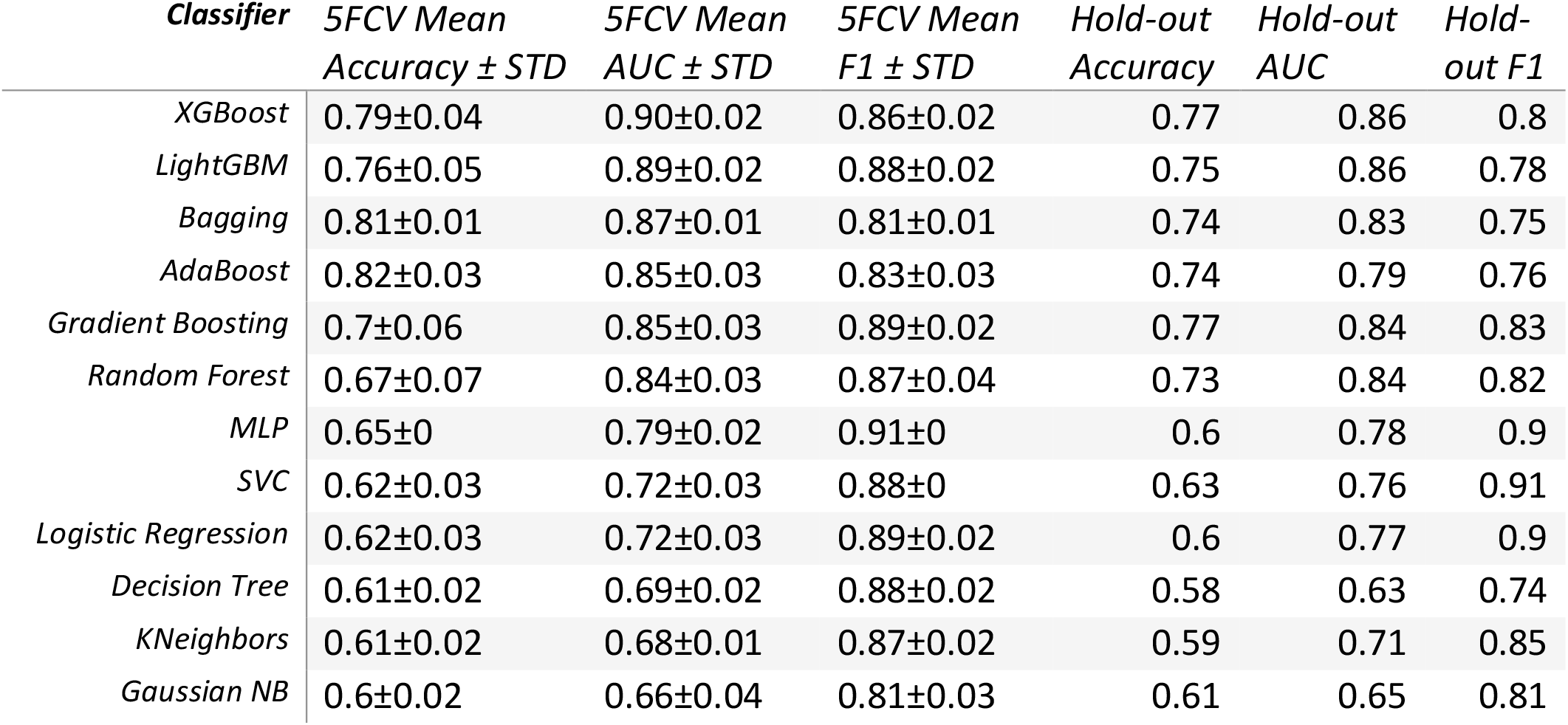
Performance summary of the 12 ML classifiers evaluated in this study.

| *Classifier* | *5FCV Mean*<br>*Accuracy ± STD* | *5FCV Mean*<br>*AUC ± STD* | *5FCV Mean*<br>*F1 ± STD* | *Hold-out*<br>*Accuracy* | *Hold-out*<br>*AUC* | *Hold-*<br>*out F1* |
| --- | --- | --- | --- | --- | --- | --- |
| *XGBoost* | 0.79±0.04 | 0.90±0.02 | 0.86±0.02 | 0.77 | 0.86 | 0.8 |
| *LightGBM* | 0.76±0.05 | 0.89±0.02 | 0.88±0.02 | 0.75 | 0.86 | 0.78 |
| *Bagging* | 0.81±0.01 | 0.87±0.01 | 0.81±0.01 | 0.74 | 0.83 | 0.75 |
| *AdaBoost* | 0.82±0.03 | 0.85±0.03 | 0.83±0.03 | 0.74 | 0.79 | 0.76 |
| *Gradient Boosting* | 0.7±0.06 | 0.85±0.03 | 0.89±0.02 | 0.77 | 0.84 | 0.83 |
| *Random Forest* | 0.67±0.07 | 0.84±0.03 | 0.87±0.04 | 0.73 | 0.84 | 0.82 |
| *MLP* | 0.65±0 | 0.79±0.02 | 0.91±0 | 0.6 | 0.78 | 0.9 |
| *SVC* | 0.62±0.03 | 0.72±0.03 | 0.88±0 | 0.63 | 0.76 | 0.91 |
| *Logistic Regression* | 0.62±0.03 | 0.72±0.03 | 0.89±0.02 | 0.6 | 0.77 | 0.9 |
| *Decision Tree* | 0.61±0.02 | 0.69±0.02 | 0.88±0.02 | 0.58 | 0.63 | 0.74 |
| *KNeighbors* | 0.61±0.02 | 0.68±0.01 | 0.87±0.02 | 0.59 | 0.71 | 0.85 |
| *Gaussian NB* | 0.6±0.02 | 0.66±0.04 | 0.81±0.03 | 0.61 | 0.65 | 0.81 |

### .2. Case Study: Generating a UPhAIR Report

To demonstrate the UPhAIR pipeline, we present a case study involving a single patient who was excluded from the training phase. The best-performing model, an XGBoost Classifier, predicted this hold-out patient to have an IDH-wildtype glioma. SHAP analysis revealed the five most influential features contributing to this prediction: MGMT promoter status as unmethylated (SHAP value: 0.0725), patient age in the 65–69 year range (SHAP: 0.0475), and three radiomics features, including first-order Total Energy of 166×10^9^ (SHAP: 0.0125), GLCM Cluster Prominence of 429,481.09 (SHAP: 0.0100), and GLRLM Long Run High Gray Level Emphasis of 2013.09 (SHAP: 0.0075). These clinical and imaging-derived features, together with a curated set of 1,010 relevant PubMed articles, were processed through the pipeline to generate the following report.

This report provides a clear, concise, and referenced explanation of the AI’s reasoning, making it readily interpretable and verifiable for a clinician.

## 4. Discussion

There are various challenges in the field of AI in medicine, such as a lack of generalisability, interpretation of outliers, and explanability. This study introduces UPhAIR, a hybrid interpretability pipeline that combines predictive machine learning, SHAP-based feature attribution, and LLM-driven narrative generation to improve the transparency of AI in medical applications. Applied to the prediction of IDH mutation status in glioma patients, UPhAIR generates clear, evidence-based reports that explain AI decisions in a clinically accessible way. By integrating SHAP’s quantitative rigor with the narrative fluency of Gemini 2.5 Pro, UPhAIR directly addresses the growing demand for interpretable AI in high-stakes domains like medicine, where clarity and trust are paramount [22].

Previous studies have leveraged radiomics features from multiparametric MRI for tasks like IDH mutation classification, but these models often lack explainability [23,24] In our study, the best model achieved an AUC of 0.90 ± 0.02 on 5-fold cross-validation using an extreme gradient boosting (XGBoost) classifier and a hold-out test AUC of 0.86, demonstrating strong predictive performance. However, the primary goal was not just accuracy but interpretability. UPhAIR addresses this need by focusing on transparent, clinically meaningful features and generating context-rich, literature-supported narratives that align with clinical reasoning. For example, a UPhAIR report might explain an IDH mutation prediction by detailing how specific radiomic characteristics and clinical variables, such as patient age or MGMT promoter status, contributed to the AI’s conclusion. By making predictions verifiable and grounded in evidence, these narratives enhance clinician trust. This transparency reduces cognitive burden and supports faster, more confident decision-making in real-world clinical environments, addressing a key barrier to AI adoption in healthcare [25].

UPhAIR uses SHAP to identify and rank key predictive features such as clinical and tumor radiomic characteristics based on Shapley values. These numerical insights are passed to an LLM that generates plain-language explanations grounded in a curated, domain-specific knowledge base of peer-reviewed literature. Unlike visualization-based XAI methods like Grad-CAM [26] or attention mechanisms [27], which lack contextual depth, UPhAIR produces narrative explanations that mirror clinical reasoning and fit seamlessly into decision-making workflows [28].

To enhance reliability and mitigate hallucinations, UPhAIR applies principles of RAG [29]. This ensures that LLM-generated reports are grounded in verified sources and aligned with the clinical context, which is an increasingly critical requirement in medical AI [30]. Structured prompting and one-shot learning further enhance the accuracy and coherence, enabling the reproducibility of the generated explanations.

UPhAIR is also model-agnostic and can be integrated with any predictive model that outputs feature importance scores. Its flexible architecture also enables adaptation across medical domains, simply by updating the underlying knowledge base. This design aligns with global calls for ethical and transparent AI in healthcare [31] and positions UPhAIR as a scalable, responsible solution for explainable clinical AI.

The example reports in Figure 2 illustrate UPhAIR’s ability to provide clear, literature-supported explanations for correctly classified predictions across both IDH-wildtype and IDH-mutant outcomes. In the IDH-wildtype case (Panel A), dominant features such as very low first-order minimum intensity (reflecting necrosis) and low GLDM Large Dependence High Gray Level Emphasis (indicating aggressive heterogeneity patterns) align with known radiomic signatures of glioblastoma-like tumors. In the IDH-mutant case (Panel B), the model appropriately weighted absent MGMT methylation (linked to G-CIMP phenotype) and low zone entropy/run length non-uniformity (indicating relative homogeneity) despite a high patient age pushing in the opposite direction. These narratives demonstrate how UPhAIR translates complex SHAP contributions into clinically interpretable insights grounded in established literature. While the current examples focus on correct predictions to highlight successful feature reasoning, the same pipeline can be applied to misclassified cases. In such scenarios, the generated reports would transparently reveal which features (e.g., outlier radiomic values or conflicting clinical variables) drove the erroneous output, enabling clinicians and developers to identify potential dataset biases, feature limitations, or cases requiring multimodal confirmation. This auditing capability further strengthens trust in AI-assisted predictions, even when the model errs, and will be explored in future validation studies with larger cohorts including error analysis.

**Figure 1.**
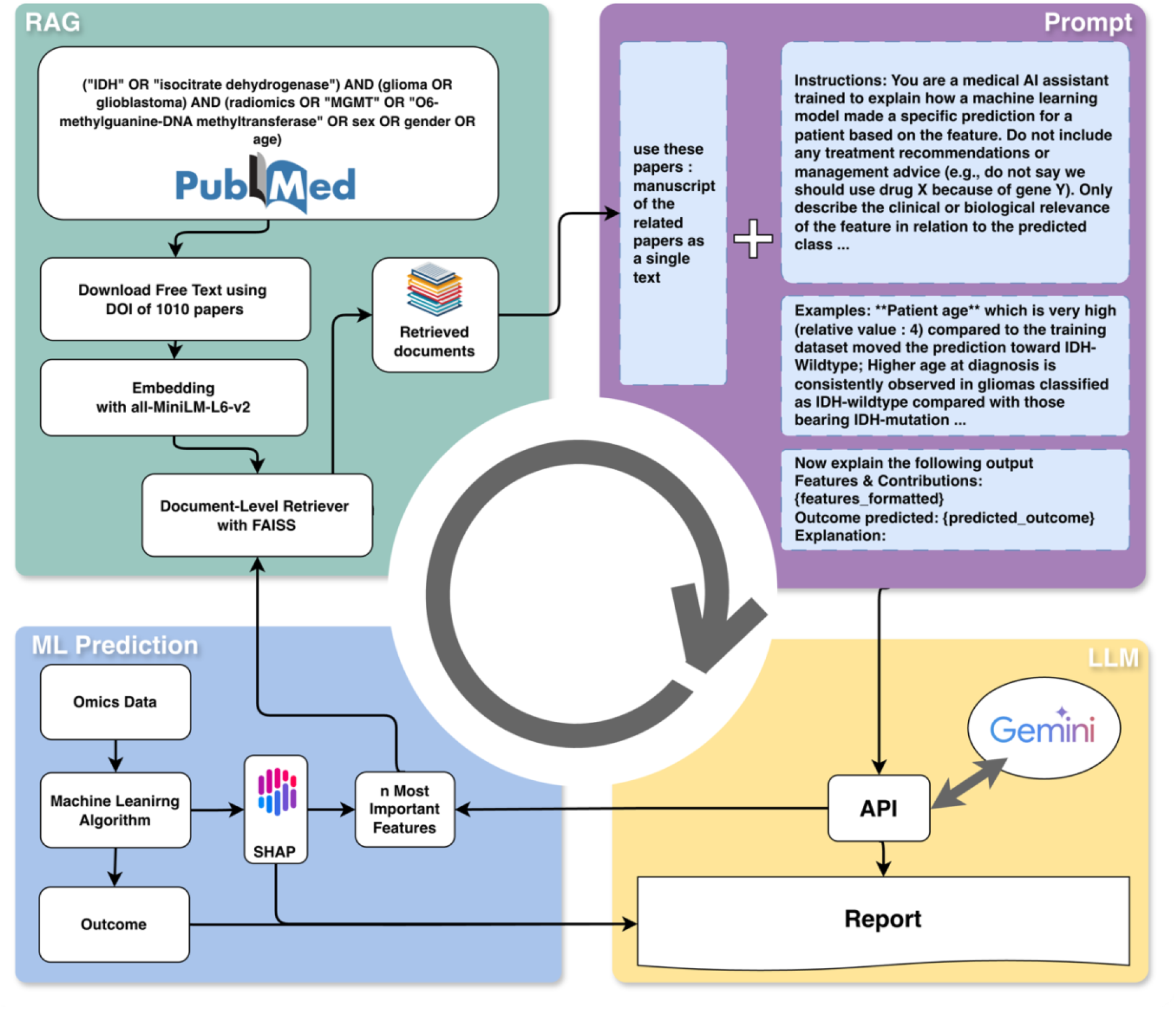
Overview of the complete UPhAIR pipeline, including retrieval-augmented generation (RAG), machine learning-based outcome prediction, and large language model (LLM)-driven report generation for IDH status prediction in glioma.

**Figure 2.**
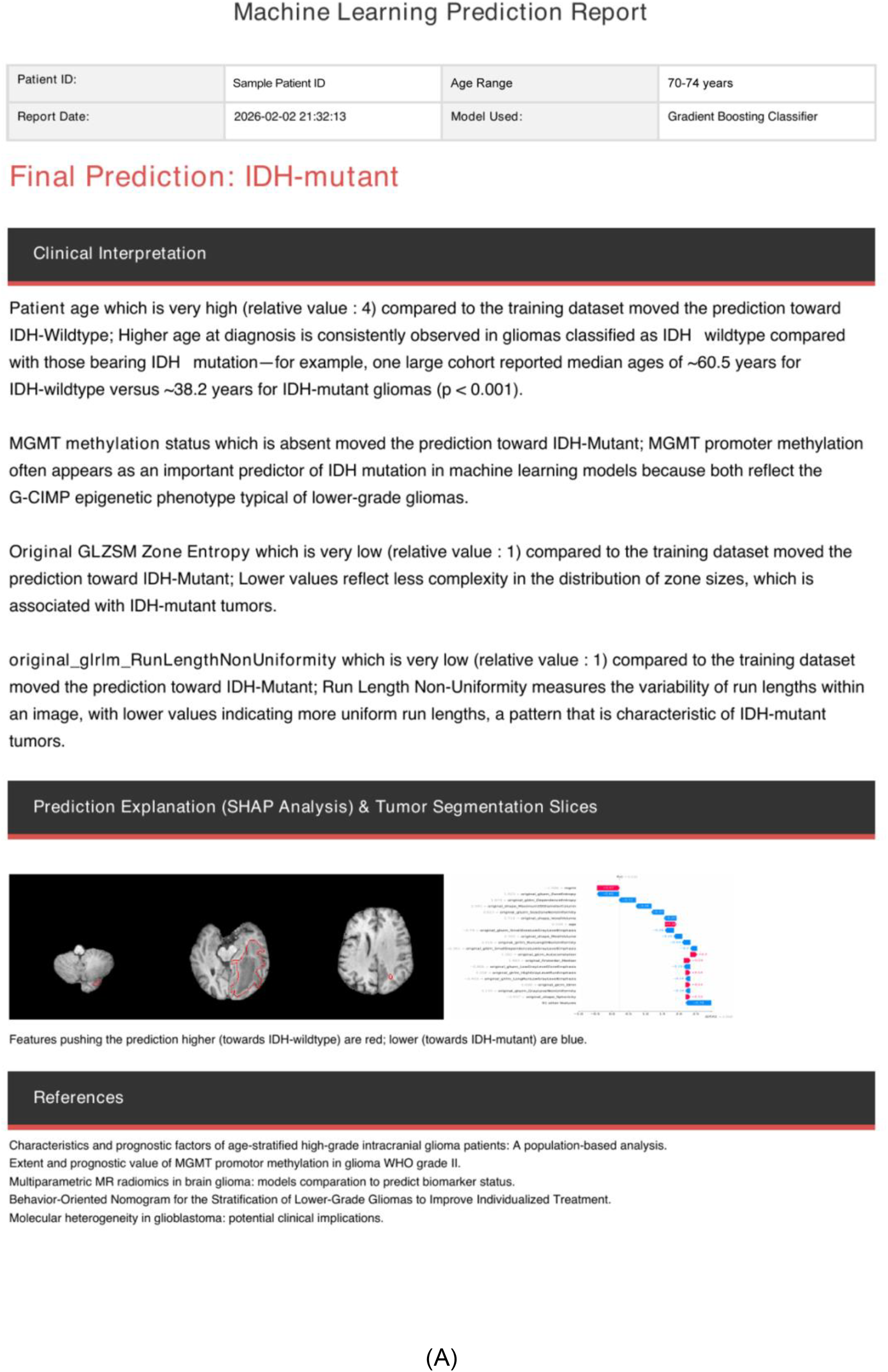
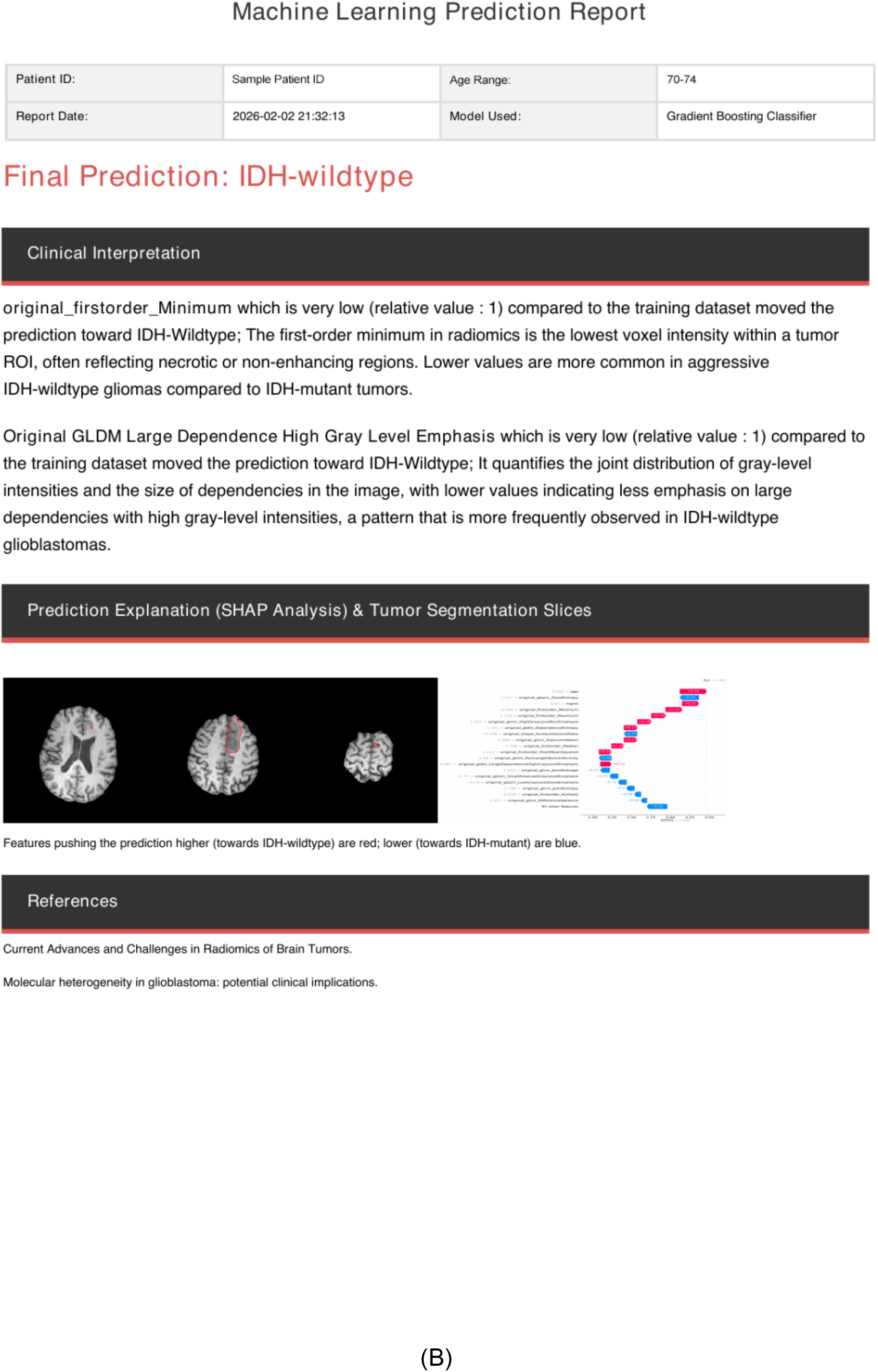
Example UPhAIR-generated post-hoc explanation reports for two correctly classified glioma cases. (A) Correctly predicted IDH-mutant glioma. The report shows patient details, prediction, SHAP analysis with segmentation, and interpretations: very high patient age (relative value: 4) pushing toward wildtype (but overridden by other features), absent MGMT methylation favoring mutant via G-CIMP phenotype, very low Original GLSZM Zone Entropy (relative value: 1) and original_glrlm_RunLengthNonUniformity (relative value: 1) reflecting lower complexity/uniformity characteristic of IDH-mutant tumors. (B) Correctly predicted IDH-wildtype glioma. The report integrates patient details, final prediction, SHAP waterfall plot with tumor segmentation slices, and clinical interpretations of key features: very low original_firstorder_Minimum (relative value: 1) reflecting necrosis/non-enhancing regions typical of aggressive IDH-wildtype tumors, and very low Original GLDM Large Dependence High Gray Level Emphasis (relative value: 1) indicating patterns more frequent in IDH-wildtype glioblastomas. Literature-grounded explanations are provided.

The effectiveness of UPhAIR’s narrative reports depends largely on the quality and completeness of its curated knowledge base. In our prediction task, we had access only to limited clinical variables, with no genomics data, and the imaging dataset was restricted to T1 sequences without T2 or FLAIR. When the underlying corpus is incomplete or biased, the generated explanations can lack clinical accuracy or relevance. The capabilities of the LLM also influence report quality; more advanced models could generate clearer and more nuanced narratives. Furthermore, the system has not yet been tested in real-world clinical environments, so its impact on clinician trust, diagnostic confidence, or decision-making is unknown. Lastly, UPhAIR’s reports are static in nature, which limits interactivity compared to dynamic or conversational systems.

Future research will focus on evaluating UPhAIR’s usability through human-computer interaction studies with clinicians and extending its application to other domains. Incorporating more advanced LLMs and implementing dynamic, continuously updated knowledge bases may further enhance the clarity and relevance of generated explanations. Additionally, developing standardized metrics to assess explanation fidelity, clinical usefulness, and decision support value will be essential to demonstrating UPhAIR’s effectiveness as a trustworthy AI tool in healthcare.

## 5. Conclusion

The UPhAIR pipeline represents a significant advancement in the field of explainable AI for medicine. By combining the analytical rigor of SHAP with the communicative power of LLMs, we have created a tool that can demystify complex AI predictions and make them accessible to clinicians. We believe that this hybrid approach has the potential to accelerate the adoption of AI in healthcare, fostering a new era of data-driven, transparent, and collaborative clinical predictions.

## Data Availability

All data produced are available online at The Cancer Imaging Archive (TCIA).

https://www.cancerimagingarchive.net/collection/upenn-gbm/

https://www.cancerimagingarchive.net/collection/ucsf-pdgm/

