## Supplemental File 1 for "UPhAIR: A Hybrid Pipeline for Generating Understandable Post-hoc AI Reports in Glioma IDH Mutation Status Prediction"

You are a medical AI assistant trained to explain how a machine learning model made a specific prediction for a patient based on the feature.
Your explanation must be clear and readable for physicians and clinicians, using clinical reasoning supported by relevant literature provided below.
Do not include any treatment recommendations or management advice (e.g., do not say we should use drug X because of gene Y).
Only describe the clinical or biological relevance of the feature in relation to the predicted class.
strictly follow the structure and tone of the examples provided.

Your instructions:

- Dont bring any additional introduction and explanation before **feature**
- Explain the model's prediction based on the given feature realtive value and their SHAP values.
- For each key feature:
    * when the outcome is IDH-mutant do not explain features with positive shap value
    * when the outcome is IDH-wildtype do not explain features with negetive shap value
    * Report its value and SHAP contribution
        (SHAP : positive value) --> moved the prediction toward IDH-wildtype
        (SHAP : negetive value) --> moved the prediction toward IDH-mutant
    * Max three sentences
    * Do not include new feature if it is against previous features.
    * Provide a clinical or biological interpretation of its role in the model's decision.
    * Do not include features that are not explainable.
    * [Relative values] indicate how the feature compares to the training cohort:
        - (Relative Value : 1) : Very Low (below 25th percentile)
        - (Relative Value : 2) : Low (25th to 50th percentile)
        - (Relative Value : 3) : High (50th to 75th percentile)
        - (Relative Value : 4) : Very High (above 75th percentile)
    * Binary features should be reported as
        "1.00" --> "Present"
        "0.00" --> "Absent"
    * Support relationships between features and {predicted_outcome_0} using evidence from below Literature.
        - All features except clinical features are derived from radiomic analysis of MRI scans.
        - if an explanation is in contrast with the litrature then return (*) as output

========================

EXAMPLE 1 where the predicted outcome is IDH-Wildtype, age value is in range of 85-90, and SHAP value is positive and Relative Value is 4:
**Patient age** which is very high (relative value : 4) compared to the training dataset moved the prediction toward IDH-Wildtype; Higher age at diagnosis is consistently observed in gliomas classified as IDH-wildtype compared with those bearing IDH-mutation—for example, one large cohort reported median ages of ~60.5 years for IDH-wildtype versus ~38.2 years for IDH-mutant gliomas (p < 0.001).

EXAMPLE 2 where the predicted outcome is IDH-Mutant, MGMT value is 0.00, and SHAP value is positive and Relative Value is binary:
**MGMT methylation** status which is absent moved the prediction toward IDH-Mutant; MGMT promoter methylation often appears as an important predictor of IDH mutation in machine learning models because both reflect the G-CIMP epigenetic phenotype typical of lower-grade gliomas.

EXAMPLE 3 where the predicted outcome is IDH-Wildtype, First order minimum value is 10, and SHAP value is positive and Relative Value is 1:
**First order minimum** which is very low (relative value : 1) compared to the training dataset moved the prediction toward IDH-Wildtype; The first-order minimum in radiomics is the lowest voxel intensity within a tumor ROI, often reflecting necrotic or non-enhancing regions. Lower values are more common in aggressive IDH-wildtype gliomas compared to IDH-mutant tumors.

EXAMPLE 4 where the predicted outcome is IDH-Mutant, Size Zone Non-Uniformity Normalized is 10 and SHAP value is positive and Relative Value is 1:
**Size Zone Non-Uniformity Normalized** which is very low (relative value : 1) compared to the training dataset moved the prediction toward IDH-Mutant; it captures the variability in the sizes of homogeneous intensity zones within a tumor, with lower values reflecting less internal heterogeneity, patterns that are characteristic of IDH-mutant tumors.

========================

**Retrieved Evidence from Literature:**
---
{retrieved_context}
---

Feature set:
{features_formatted}

**Supplemental Figure 1:** Complete system prompt used by the large language model in the UPhAIR pipeline. This figure presents the full, structured prompt fed to the LLM (Google Gemini 2.5 Pro) for generating clinically readable, evidence-based explanations of machine learning predictions for IDH mutation status in glioma patients. The prompt enforces a strict format (feature-by-feature explanations, SHAP value interpretation, relative value categories, binary feature wording, maximum three sentences per feature), restricts explanations to features supporting the predicted class, and requires grounding in provided literature excerpts. Example outputs are included for both IDH-wildtype and IDH-mutant predictions to illustrate tone, structure, and adherence to clinical reasoning without treatment recommendations.

| *Center* | | *UPENN* | *UCSF* |
|---|---|---|---|
| *Age (mean)* | | 62.9 | 56.7 |
| *Sex* | Male | 194 | 366 |
| | Female | 244 | 292 |
| *MGMT* | Methylated | 138 | 296 |
| | Unmethylated | 151 | 113 |
| | Unknown | 321 | 77 |
| *IDH* | Mutant | 112 | 103 |
| | Wildtype | 498 | 383 |

**Supplemental Table 1:** Patient demographics from the UPENN and UCSF cohorts used in model training and evaluation. This table summarizes key demographic and molecular characteristics of the glioma patients across two centers (UPENN and UCSF). Columns include mean age, sex distribution (male/female counts), MGMT promoter methylation status (methylated, unmethylated, unknown), and IDH mutation status (mutant/wildtype counts). These variables were incorporated as clinical features alongside MRI-derived radiomics in the prediction of IDH status.

| model | mcc_threshold | cv_bal_acc_mean | cv_mcc_mean | cv_auroc_mean | cv_f1_mean | cv_brier_mean | cv_bal_acc_std | cv_mcc_std | cv_auroc_std | cv_f1_std | cv_brier_std | test_bal_acc | test_mcc | test_auroc | test_f1 | test_brier |
|---|---|---|---|---|---|---|---|---|---|---|---|---|---|---|---|---|
| AdaBoostClassifier | 0.45 | 0.82 | 0.53 | 0.85 | 0.83 | 0.18 | 0.03 | 0.06 | 0.03 | 0.03 | 0.01 | 0.74 | 0.39 | 0.79 | 0.76 | 0.2 |
| Bagging | 0.75 | 0.81 | 0.5 | 0.87 | 0.81 | 0.11 | 0.01 | 0.02 | 0.01 | 0.01 | 0 | 0.74 | 0.39 | 0.83 | 0.75 | 0.14 |
| XGBoost | 0.9 | 0.79 | 0.51 | 0.89 | 0.86 | 0.1 | 0.04 | 0.03 | 0.02 | 0.02 | 0 | 0.77 | 0.43 | 0.86 | 0.8 | 0.1 |
| LightGBM | 0.8 | 0.76 | 0.49 | 0.89 | 0.88 | 0.09 | 0.05 | 0.05 | 0.02 | 0.02 | 0 | 0.75 | 0.41 | 0.86 | 0.78 | 0.1 |
| Gradient Boosting | 0.9 | 0.7 | 0.45 | 0.85 | 0.89 | 0.11 | 0.06 | 0.05 | 0.03 | 0.02 | 0 | 0.77 | 0.44 | 0.84 | 0.83 | 0.1 |
| RandomForestClassifier | 0.6 | 0.67 | 0.4 | 0.84 | 0.87 | 0.13 | 0.07 | 0.07 | 0.03 | 0.04 | 0.01 | 0.73 | 0.38 | 0.84 | 0.82 | 0.13 |
| MLP | 0.1 | 0.65 | 0.43 | 0.79 | 0.91 | 0.14 | 0 | 0.01 | 0.02 | 0 | 0.01 | 0.6 | 0.36 | 0.78 | 0.9 | 0.16 |
| SVC_rbf | 0.2 | 0.62 | 0.3 | 0.72 | 0.88 | 0.19 | 0.03 | 0.05 | 0.03 | 0 | 0.01 | 0.63 | 0.43 | 0.76 | 0.91 | 0.17 |
| Logistic Regression | 0.15 | 0.62 | 0.34 | 0.72 | 0.89 | 0.2 | 0.03 | 0.09 | 0.03 | 0.02 | 0.01 | 0.6 | 0.38 | 0.77 | 0.9 | 0.17 |

| | | | | | | | | | | | | | | | |
|---|---|---|---|---|---|---|---|---|---|---|---|---|---|---|---|
| DecisionTreeClassifier | 0.4 | 0.61 | 0.29 | 0.69 | 0.88 | 0.21 | 0.02 | 0.02 | 0.02 | 0.02 | 0.01 | 0.58 | 0.14 | 0.63 | 0.74 | 0.25 |
| KNeighborsClassifier | 0.2 | 0.61 | 0.27 | 0.68 | 0.87 | 0.26 | 0.02 | 0.04 | 0.01 | 0.02 | 0.01 | 0.59 | 0.19 | 0.71 | 0.85 | 0.26 |
| GaussianNB | 0.05 | 0.6 | 0.19 | 0.66 | 0.81 | 0.27 | 0.02 | 0.02 | 0.04 | 0.03 | 0.01 | 0.61 | 0.21 | 0.65 | 0.81 | 0.3 |

**Supplemental Table 2:** Cross-validation and test-set performance metrics for all 12 evaluated classifiers. This table reports comprehensive performance results for the classifiers trained to predict IDH mutation status (IDH-mutant vs. IDH-wildtype) using radiomics and clinical features. Metrics include the MCC (Matthews correlation coefficient) threshold used, 5-fold cross-validation means and standard deviations for balanced accuracy, MCC, AUROC, F1 score, and Brier score, as well as final test-set values. The table highlights XGBoost as one of the top performers (test AUROC 0.86, balanced accuracy 0.77), guiding model selection for the UPhAIR pipeline.

| | |
|---|---|
| AdaBoostClassifier | {'clf__algorithm': 'SAMME', 'clf__learning_rate': 0.1, 'clf__n_estimators': 50} |
| Bagging | {'clf__n_estimators': 50} |
| XGBoost | {'clf__colsample_bytree': 1, 'clf__eval_metric': 'logloss', 'clf__learning_rate': 0.1, 'clf__max_depth': 10, 'clf__n_estimators': 50, 'clf__subsample': 1, 'clf__use_label_encoder': False} |
| LightGBM | {'clf__boosting_type': 'dart', 'clf__learning_rate': 0.1, 'clf__max_depth': 5, 'clf__n_estimators': 100, 'clf__num_leaves': 15} |
| GradientBoosting | {'clf__max_depth': 5, 'clf__max_features': 'sqrt', 'clf__min_samples_leaf': 4, 'clf__min_samples_split': 2} |
| RandomForestClassifier | {'clf__max_depth': 10, 'clf__max_features': 'sqrt', 'clf__min_samples_leaf': 4, 'clf__min_samples_split': 5} |
| MLP | {'clf__activation': 'relu', 'clf__learning_rate': 'invscaling', 'clf__max_iter': 100} |
| SVC_rbf | {'clf__C': 0.1, 'clf__degree': 3, 'clf__kernel': 'linear', 'clf__probability': True} |
| LogisticRegression | {'clf__C': 10, 'clf__penalty': 'l2', 'clf__solver': 'sag'} |
| DecisionTreeClassifier | {'clf__max_depth': 10, 'clf__max_features': 'sqrt', 'clf__min_samples_leaf': 4, 'clf__min_samples_split': 5} |
| KNeighborsClassifier | {'clf__algorithm': 'auto', 'clf__n_neighbors': 9, 'clf__weights': 'distance'} |
| GaussianNB | {'clf__var_smoothing': 1e-06} |

**Supplemental Table 3:** Optimized hyperparameters for each of the 12 classifiers after tuning. This table lists the final hyperparameter configurations selected for each classifier (e.g., AdaBoost: algorithm='SAMME', learning_rate=0.1, n_estimators=50; XGBoost: learning_rate=0.1, max_depth=10, n_estimators=50, etc.). Hyperparameters were determined via grid search or algorithm-specific optimization to maximize predictive performance on the glioma IDH classification task. These settings correspond to the performance metrics reported in Supplemental Table 2.
